# Heterozygous and Homozygous *RFC1 AAGGG* Repeat Expansions are Common in Idiopathic Peripheral Neuropathy

**DOI:** 10.1101/2025.04.18.25325809

**Authors:** Zitian Tang, Sinem S. Ovunc, Ryo Iwase, Elle Mehinovic, Simone Thomas, Jenna Ulibarri, Zefan Li, Dustin Baldridge, Carlos Cruchaga, Matt Johnson, Jeffrey Milbrandt, Brian Callaghan, PNRR Study Group, Ahmet Höke, Peter K. Todd, Sheng Chih Jin

**Affiliations:** Department of Genetics, Washington University School of Medicine, St. Louis, MO, USA; Department of Neurology, University of Michigan, Ann Arbor, MI, USA; Department of Neurology, Johns Hopkins University School of Medicine, Baltimore, MD, USA; Department of Pediatrics, Washington University School of Medicine, St. Louis, MO, USA; NeuroGenomics and Informatics Center, Washington University School of Medicine in St. Louis, St. Louis, MO, USA; Needleman Center for Neuro-metabolism and Axonal Therapeutics, St. Louis, MO, USA; McDonnell Genome Institute, Washington University School of Medicine, St. Louis, MO, USA; Veterans Affairs Medical Center, Ann Arbor, MI, USA

## Abstract

**Objective:** Biallelic intronic AAGGG repeat expansions in *RFC1* cause Cerebellar Ataxia with Neuropathy and Vestibular Areflexia Syndrome and may also contribute to isolated sensory neuropathy. The clinical significance of both heterozygous and biallelic *RFC1* expansions in more diverse patient populations remains unclear—partly due to the absence of accurate, user-friendly computational pipelines specifically tailored for tandem repeat analysis.

**Methods:** To discern the relationship between *RFC1* expansions and idiopathic peripheral neuropathy (iPN), we performed whole-genome sequencing (WGS) followed by PCR-based confirmation in a large, well-characterized U.S. cohort consisting of 788 iPN patients (369 pure small fiber neuropathy (SFN), 266 sensorimotor, 144 pure sensory, and 9 pure motor). We developed an integrative pipeline combining ExpansionHunter Denovo and ExpansionHunter coupled with unsupervised clustering to reliably detect and genotype *RFC1* expansions from short-read WGS data, achieving 98.2% concordance with repeat-primed PCR based validation.

**Results:** Biallelic *RFC1* expansions were present in only one out of 854 controls but present in 2.3% of iPN patients (Fisher’s exact *p* = 2×10^-5^), including 6.9% of pure sensory, 1.1% of SFN, and 1.5% of sensorimotor neuropathy, indicating that motor nerve involvement should not exclude patients from *RFC1* repeat screening. We also observed an increased frequency of monoallelic expansions in iPN compared to controls (9.4% versus 6.3%; Fisher’s exact *p* = 0.02), without evidence of secondary mutations or expansions on the other allele.

**Interpretation:** Our approach provides a robust, cost-effective method for detecting *RFC1* expansions from WGS data. Our findings indicate that both heterozygous and homozygous AAGGG repeat expansions in *RFC1* contribute to development of iPN.

## Introduction

Peripheral neuropathy (PN) is among the most common neurological disorders, with an estimated prevalence of 12–20% in the United States and up to 30% in older adults.^1^ PN causes a range of debilitating symptoms including pain, sensory loss, imbalance, and weakness. These manifestations, although rarely lethal, contribute to significant humanistic and socioeconomic burden, reflected in patients’ reduced quality of life and severe impairments in daily functioning.^2^ Approximately one-third of all PN cases are classified as idiopathic (iPN), wherein no definitive etiology can be determined despite thorough and appropriate clinical evaluation.^3^ As upwards of 18% of late-onset iPN may be explained by genetic causes,^4^ neurology clinics increasingly implement gene-panel based testing for potential pathogenic single nucleotide variants (SNVs) and copy number variations (CNVs).^5^ Despite these efforts, most iPN patients remain undiagnosed even after gene panel testing.

Short tandem repeats (STRs), defined as repetitive units of 1 to 6 base pairs (bp) adjacent to each other, constitute around 8% of the human genome.^6–8^ Due to replication slippage and mismatch repair errors that occur during DNA replication and RNA transcription, STRs have among the highest mutation rates of all genetic variant classes.^6,9,10^ Genomic instability caused by pathogenic STR expansion contributes to over 70 human disorders, many of which are neurological.^11^ Recent large-scale initiatives such as the 1000 Genomes Project^12^ and the Simons Genome Diversity Project^13^ have cataloged extensive polymorphic STRs in healthy individuals, revealing that most loci exhibit multiple common alleles and that each genome carries thousands of STR variants relative to the reference. The increasing accuracy of short-read, high-coverage whole-genome sequencing (WGS), coupled with specialized bioinformatic tools, has further refined our understanding of the prevalence and impact of STR expansions on human health.^12,13^ Nonetheless, current STR detection software—such as ExpansionHunter Denovo (EHdn)^14^, ExpansionHunter (EH)^15^, GangSTR^16^, and exSTRa^17^— faces several limitations, including reliance on predefined repeat catalogs, incomplete assessment of novel motifs, limited ability to capture zygosity, and complex computational requirements that pose hurdles for routine clinical use. Recent work has begun to assess the accuracy of *RFC1* expansion genotyping from short-read WGS. ^18^

STR expansions have emerged as a key contributor to PN pathogenesis. In 2019, a biallelic non-reference AAGGG repeat expansion in the second intron of the *RFC1* (Replication Factor C Subunit 1) gene was identified as the most common cause of cerebellar ataxia, neuropathy, and vestibular areflexia syndrome (CANVAS), a multisystemic disorder characterized by progressive imbalance, sensory neuronopathy and cerebellar atrophy.^19–21^ *RFC1* encodes the principal subunit of the replication factor C complex, essential for DNA polymerases and repair through ATP-dependent loading of proliferating cell nuclear antigen onto DNA strands. ^22,23^ Despite its ubiquitous expression and critical role in genomic integrity maintenance, particularly in DNA damage recognition and repair pathways, the pathological mechanism linking *RFC1* expansions to disease remains unclear.^22,23^ Pathogenic alleles typically involve replacement of the wild-type (AAAAG)_11_ repeat motif with over 250 repeats of AAGGG.^20,24^ Since the original discovery, additional pathogenic repeat motifs, including ACAGG, AAGGC, AGGGC, and AGAGG, have been identified either in the homozygous or compound heterozygous state with the AAGGG expansion.^25,26^ It is now recognized that AAAGG repeats, initially considered non-pathogenic, may cause disease at substantially expanded repeat lengths.^26^ The identification of rare compound heterozygous cases with *RFC1* truncating or splicing mutations instead of wild-type (AAAAG)_11_ repeat alongside a single-allele expansion aligns with the recessive inheritance pattern of CANVAS, and suggest a potential loss-of-function mechanism.^27–29^ However, to date, no alterations in *RFC1* expression, splicing, or DNA repair function have been correlated with the repeat expansions; thus, the precise mechanism by which the *RFC1* repeat expansions causes disease remains unknown.^19,20,29,30^

More recently, this same biallelic expansion was also observed in an Italian patient cohort with chronic idiopathic axonal polyneuropathy.^19–21^ These studies suggested biallelic AAGGG repeat expansions as a causative phenomenon isolated to sensory neuropathy, but absent in those with sensory-motor neuropathy. Nevertheless, whether the results of this standalone study are representative of the impact of AAGGG repeats on susceptibility to PN pathogenesis within the broader population, and whether repeats in the heterozygosity state confer additional risk for neuropathy, remains largely unexplored.

Here we describe analysis of STR expansions in a well-characterized U.S. iPN cohort (the Peripheral Neuropathy Research Registry, or PNRR), coupling short-read WGS with confirmatory repeat-primed PCR (RP-PCR). We developed and applied a novel automated pipeline that integrates EHdn and EH to improve the sensitivity and specificity of STR detection. Additionally, we employed unsupervised clustering to robustly classify *RFC1* genotypes to assess the impact of biallelic AAGGG expansions and the potential risk associated with monoallelic expansion carriers. By benchmarking our pipeline’s performance against repeat-primed PCR results — the current clinical gold standard for *RFC1* repeat expansion testing in the U.S. — we aim to not only to validate *RFC1* STR motif detection in the iPN population but also pave the way for broader clinical application of STR-based diagnostics in neuropathy and other neurological disorders.

## Methods

### iPN Patient Cohort

DNA samples (whole blood, buffy coat, or saliva) were obtained from 816 patients with iPN enrolled in the PNRR at the Johns Hopkins Hospital. The McDonnell Genome Institute at Washington University School of Medicine generated short-read WGS data at 30X coverage for all samples. We conducted extensive quality control (QC), excluding 18 samples that failed either sequencing or alignment and removing 10 samples not sent for PCR analysis, resulting in a final cohort of 788 iPN cases (**Table 1**). We calculated read length, number of sequencing reads, insert size, percentage reads or bases mapped, duplicate rate, and mean error rate based on results from samtools stats (samtools version 1.10),^31^ and the mean coverage from samtools depth. The post-alignment QC metrics are shown in **Table S1**.

**Table 1.**
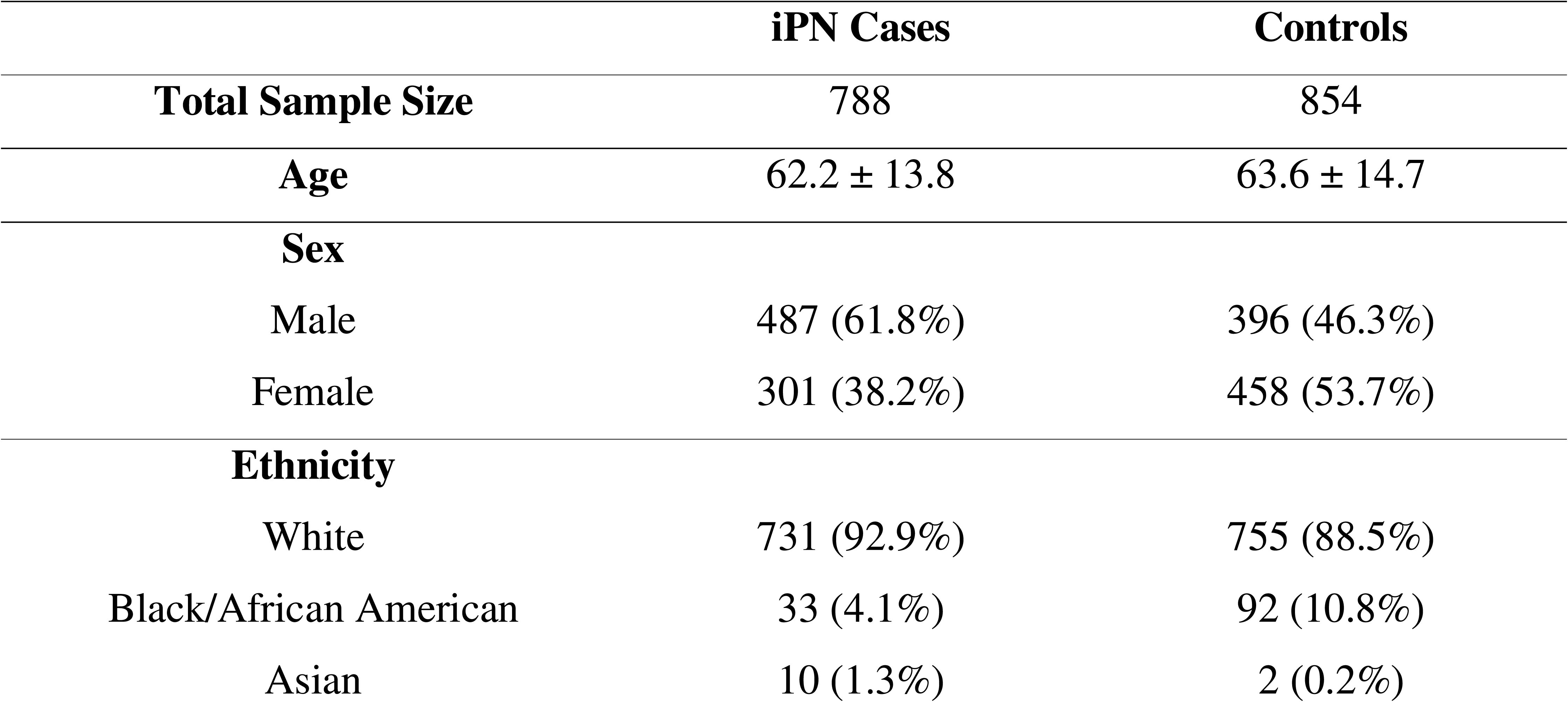
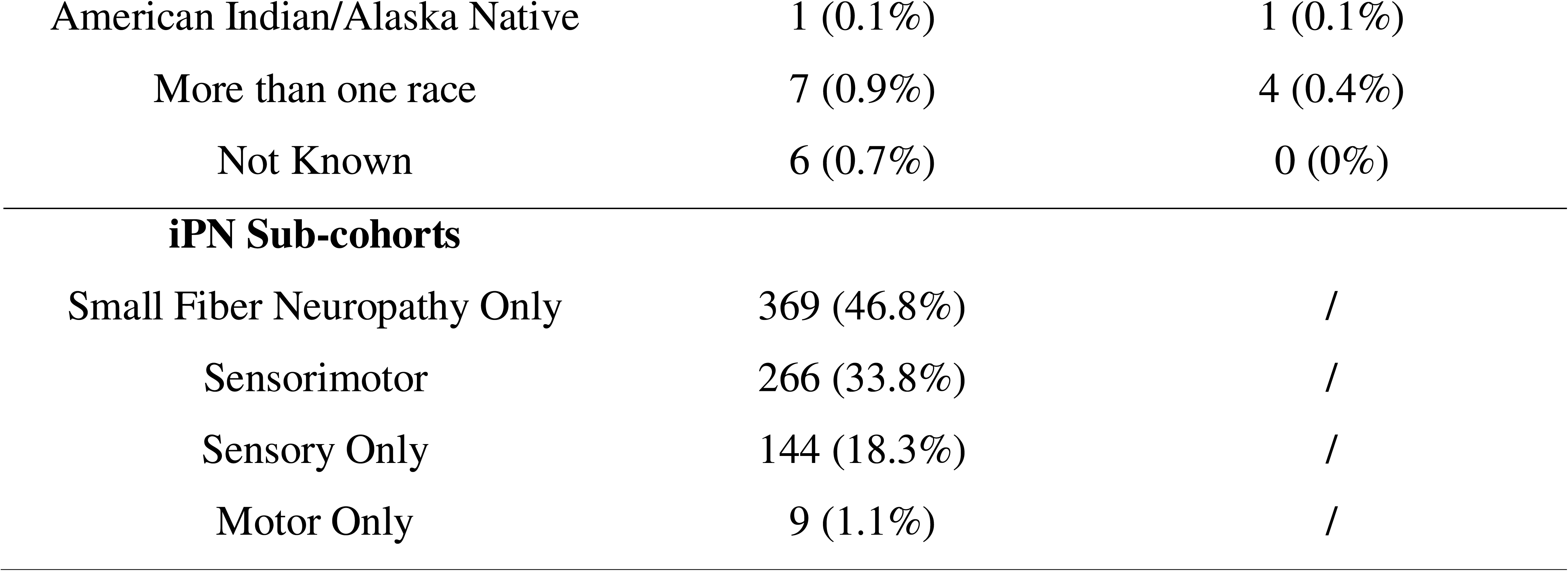
Demographic and clinical characteristics of study participants. A total of 788 iPN cases and 854 controls were included. Demographic data show comparable age and sex distribution between iPN cases and controls. iPN cases were predominantly classified into four sub-cohorts: small fiber neuropathy only (46.8%), sensorimotor (33.8%), sensory only (18.3%), and motor only (1.1%). iPN, idiopathic peripheral neuropathy.

|  | iPN Cases | Controls |
| --- | --- | --- |
| Total Sample Size | 788 | 854 |
| Age | 62.2 ± 13.8 | 63.6 ± 14.7 |
| Sex |  |  |
| Male | 487 (61.8%) | 396 (46.3%) |
| Female | 301 (38.2%) | 458 (53.7%) |
| Ethnicity |  |  |
| White | 731 (92.9%) | 755 (88.5%) |
| Black/African American | 33 (4.1%) | 92 (10.8%) |
| Asian | 10 (1.3%) | 2 (0.2%) |
| American Indian/Alaska Native | 1 (0.1%) | 1 (0.1%) |
| More than one race | 7 (0.9%) | 4 (0.4%) |
| Not Known | 6 (0.7%) | 0 (0%) |
| **iPN Sub-cohorts** |  |  |
| Small Fiber Neuropathy Only | 369 (46.8%) | / |
| Sensorimotor | 266 (33.8%) | / |
| Sensory Only | 144 (18.3%) | / |
| Motor Only | 9 (1.1%) | / |

The demographic data for the iPN cohort are detailed in **Table 1**, with an average age at visit of 62.2 years old (standard deviation [SD] = 13.6), a sex ratio slightly skewed toward male (61.8%) over female (38.2%), and a majority White ethnic composition (92.4%). The institutional review boards of Washington University School of Medicine (Protocol no. 202401194), Johns Hopkins University School of Medicine (Protocol no. NA_00047435), and University of Michigan (Protocol no. HUM00244538) approved the protocols.

### iPN Categorization

For comparison purposes, we classified the iPN cohort into four subcategories. The sensory iPN subcategory (N = 144) includes patients with large and small fiber sensory involvement based on electrophysiology and neurological examination. The SFN subcategory (N = 369) comprises patients with involvement of only small sensory fibers as determined by normal nerve conduction studies but with an abnormal skin biopsy and/or neurological examination.

The sensorimotor iPN subcategory (N = 266) consists of patients with a sensorimotor involvement as determined by electrophysiology and neurological examination. Finally, the sensory and SFN subcategory (N = 513) includes patients with either sensory iPN or SFN (**Table 1**).

### Control Cohort

To effectively filter out STRs present in the general population, we incorporated a control cohort matched for ethnic background and age distribution. The control cohort (N = 854) comprises 464 cognitively normal elderly individuals enrolled as controls in the Knight Alzheimer’s Disease Research Center (ADRC) at Washington University and 390 unaffected parents aged 40 years or older from the Undiagnosed Mendelian Disease (UMD) study at Washington University. All PCR-free short-read WGS for controls were generated at Washington University on Illumina NovaSeq platforms. Overall, the control cohort closely resembles the iPN cohort in mean age (63.6 ± 14.7 years) and racial composition (88.5% White), as well as in sequencing QC metrics (**Table 1** and **S1**).

### Novel STR Expansion Detection Pipeline based on WGS

Initial STR expansion detection was performed using ExpansionHunter Denovo (EHdn)^14^ and ExpansionHunter (EH)^15^ on BAM files generated from patient sample sequencing (**Figure 1A**). These data were aligned to the GRCh38 reference genome using Parabricks *pbrun germline* pipeline (version 4.0.0-1)^32^, which utilized BWA-MEM^33^ and STAR^34^ as the sequencing aligners. We then used EHdn (v0.9.1) to analyze the aligned BAM files and performed genome-wide STR scanning without reliance on predefined catalogs. We utilized ANNOVAR^35^ to annotate genes where STRs located, and the RepeatMasker database^36^ to annotate known gene-motif pairs. Next, we prioritized motifs detected in the *RFC1* gene and created a *RFC1* STR catalog using gene information, repeat motifs, and genomic coordinates from EHdn output. We provided this catalog alongside the BAM files to EH (v5.0.0) for further refinement of our STR detection. EH acts not only as a genotyping tool but also an *in silico* validation of EHdn calls. In the final analysis, we retained only STR motifs detected by both tools, requiring at least 10% of carriers detected by both tools. For each motif, we then defined carriers as the union of samples identified by EHdn and those with at least one allele labeled with “INREPEAT” by EH.

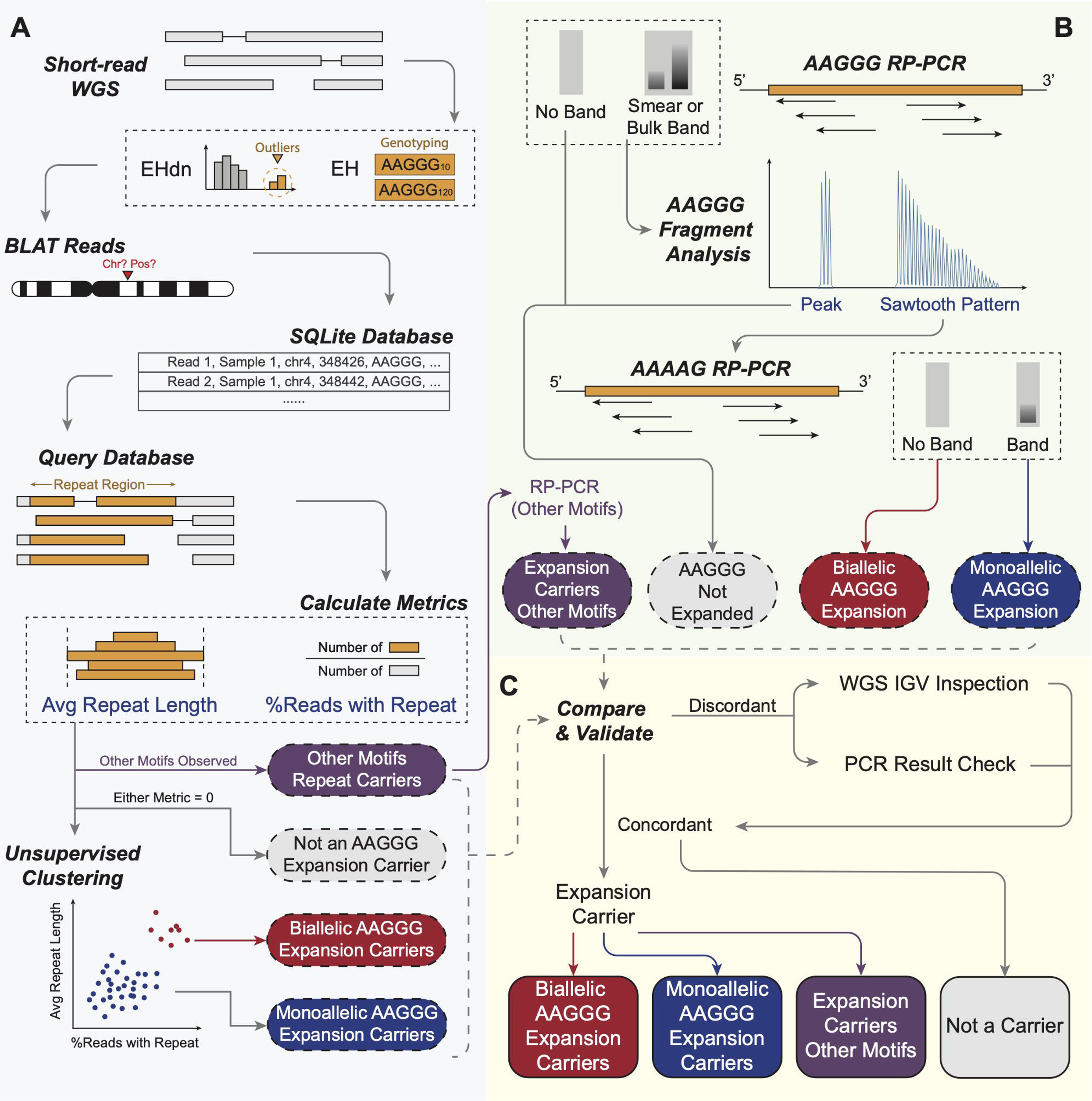

### STR Motif Validation and Carriers Identification

To validate the STR motifs identified in the previous step, we generated SAM files for each carrier in the potential STR expansion regions (**Figure 1A**). Here, we defined the repeat window (i.e., region-of-interest, or ROI) as 90 bp upstream and downstream from the *RFC1* repeat region in the reference genome. We initialized an SQLite database (Python 3.6.3, SQLite3 2.6.0) containing all data from the SAM files to enable precise and efficient information retrieval and querying in subsequent steps. BLAT (version v.36) was run on every read within the SAM files, generating one PSL file per carrier per STR motif. We then used an in-house Python script to calculate the top three BLAT matches and their corresponding scores, which were integrated as a new column into the database.

We identified eligible carriers for a specific STR motif by extracting all unique samples listed in the database. To quantify STR length, we determined the total length of each read from its CIGAR string and identified the longest consecutive stretch of the queried motif, allowing one mismatch per motif unit through a Hamming distance approach. All rotational variants of the motif (e.g., ACG, CGA, GAC) were treated as the same motif. The algorithm also reports other motifs recognized that might interrupt the queried motif or reside on the other allele. RP-PCR-validated alternate motif expansion carriers are listed in **Table S4**.

We next compared each read’s STR length to a predetermined threshold (defaulted to be 10% of the read length). The proportion of candidate reads, carrying repeats longer than this threshold, relative to the total read count and multiplied by 100, provides the percentage of STR-containing reads for each sample (equation 1). On the other hand, we calculated the mean STR length by dividing the sum of STR lengths in all reads by the total read count (equation 2). Samples with either a zero percentage of STR-containing reads or a zero mean STR length were classified as non-carriers in the short-read WGS analysis.

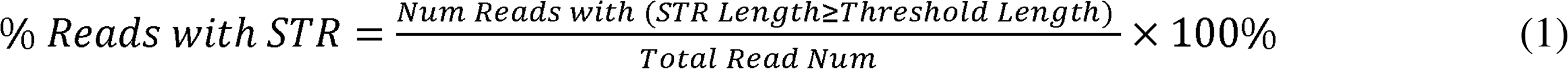

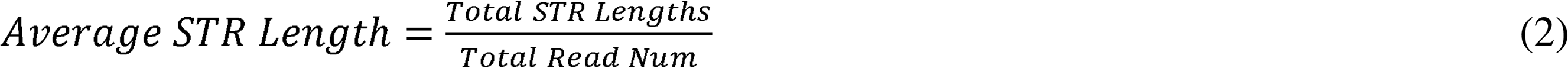

### Genotype Determination for STR Carriers

To distinguish between potential biallelic and monoallelic STR expansions among carriers, we conducted three clustering approaches based on “% Reads with STR” and “Average STR Length”, as defined by equations (1) and (2) above. Before clustering, we standardized both features using Z-score normalization to ensure comparable scales.

We determined the optimal number of clusters (K) using both the elbow and Silhouette methods (R version 4.4.1, NbClust 3.0.1) (**Figure S1A**). We then applied three clustering algorithms to identify potential biallelic and monoallelic carriers: (1) k-means with multiple 25 random starts to ensure stable cluster assignments; (2) hierarchical clustering using Ward’s method (ward.D2) with Euclidean distance that minimizes within-cluster variance; and (3) Gaussian Mixture Models using the Mclust algorithm (mclust 6.1.1). An example clustering result is shown in **Figure S1B**. Any sample identified as biallelic by at least one clustering method was considered biallelic, and all remaining samples were classified as monoallelic carriers. In cases of inconsistent results between random seeds or methods, we performed manual review using the Integrative Genomics Viewer (IGV) to confirm biallelic or monoallelic status (**Figure 1A**).

For comparison and benchmarking purposes, we also determined the genotypes for individuals detected as carriers by ExpansionHunter. As previously described, we only keep EH carriers with at least one allele’s length greater than 11 copies and was estimated from in-repeat reads (INREPEAT). Among these carriers, we further classified samples with alleles’ lengths estimated from in-repeat or flanking reads (INREPEAT/INREPEAT and INREPEAT/FLANKING) as biallelic carriers, and those from spanning reads (INREPEAT/SPANNING) as monoallelic carriers.

### Post-Analysis Case and Control Carriers Phenotype Confirmation

For cases carrying *RFC1* AAGGG expansion, we checked the iPN subtype, disease history, and other neurologically relevant test results to confirm their clinical phenotypes (**Table 2**). For control analysis, the pipeline initially detected 32 control samples as *RFC1* STR expansion carriers. We obtained the clinical phenotyping information for these carriers and excluded 10 with conditions associated with peripheral neuropathy, such as diabetes, autoimmune disorders, and neurodegenerative conditions, resulting in 22 final control carriers. The exclusion criteria for controls closely resembled that for cases, as described previously in **Methods**.

**Table 2.** Clinical characteristics of confirmed biallelic and monoallelic *RFC1* AAGGG carriers versus non-carriers in iPN cases. Chi-squared tests were conducted for sex, age at visit, painful status, and percentage carriers in each subtype across all groups. Pure sensory iPN was significantly more prevalent in biallelic carriers (56%) compared to monoallelic carriers (13%) and non-carriers (18%) (*p* < 0.001), while age at visit, sex distribution, and painful neuropathy prevalence were similar across groups (non-significant, NS). iPN, idiopathic peripheral neuropathy.

|  | **Biallelic**<br>**(N = 18)** | **Monoallelic**<br>**(N = 62)** | **Non-Carriers**<br>**(N = 708)** | **P-value** |
| --- | --- | --- | --- | --- |
| **Male** | 9 (50%) | 36 (58%) | 424 (60%) | NS |
| **Age at Visit** | 65 ± 10 | 62 ± 14 | 62 ± 14 | NS |
| **Painful Neuropathy** | 14 (78%) | 44 (71%) | 490 (70%) | NS |
| **Sensory Neuropathy** | 10 (56%) | 8 (13%) | 126 (18%) | <0.001 |
| **Sensorimotor** | 4 (22%) | 20 (32%) | 242 (3%) | NS |
| **Small Fiber Neuropathy** | 4 (22%) | 33 (53%) | 332 (47%) | NS |
| **Motor Neuropathy** | 0 (0%) | 1 (2%) | 8 (1%) | NS |

### Repeat-primed PCR

Repeat-primed PCR (RP-PCR) targeting the repeat locus was performed as previously described to identify carriers of the AAGGG expansion.^19–21^ Samples exhibiting a smear or distinct band on the gel underwent further confirmation via capillary electrophoresis conducted by Genewiz Inc. The resulting data files were analyzed using the Peak Scanner Fragment Analysis Software. Fragment analysis was subsequently employed to impute expansion lengths. Long AAGGG expansions were inferred by the presence of distinct sawtooth patterns, whereas short AAGGG repeats displayed a limited number of peaks without evidence of an extended sawtooth pattern (**Figure 1B**).

Cases with a positive RP-PCR result for the AAGGG expansion were subsequently assessed for the wild-type allele status through AAAAG RP-PCR. Those showing bands in the AAAAG RP-PCR assay were classified as likely monoallelic carriers, while those without bands were classified as likely biallelic carriers. Flanking PCR was also conducted on these cases to assess for the presence or absence of biallelic expanded repeats. To clarify cases with additional repeat motifs observed in WGS data, we performed motif-specific RP-PCR for AAAGG, ACAAG, AACGG, AAGAG, and AAAGGG, and fragment analysis to clarify their expansion status. Detailed information on primer sequences and thermocycling conditions for all PCR experiments is provided in **Table S3**.

### Benchmarking of WGS results against RP-PCR

We systematically compared WGS analysis results to those obtained via RP-PCR methods (**Figure 1C**). The WGS pipeline included intermediate EH outputs as well as final calls generated by our computational pipeline, all of which were compared with PCR data. For samples identified as carriers by only PCR or WGS pipeline, we re-examined the PCR gel electrophoresis and fragment analysis results and conducted additional inspection of the short-read WGS data using IGV. During IGV inspection, samples containing at least one correctly-aligned read—confirmed by directly BLATing the read within IGV—and exhibiting more than five consecutive repeat motifs were classified as carriers. Carriers missed by detection pipeline but rescued by IGV inspection were labeled accordingly in **Table S2**.

In addition, we compared the genotyping results from WGS analysis and the PCR method. Samples consistently classified as either biallelic or monoallelic carriers were assigned a final genotype directly, whereas samples with discordant results underwent additional IGV inspection and PCR verification. Due to the intrinsic limitations associated with short-read WGS and RP-PCR, some samples continued to show discrepant carrier status or genotype between the two methods. In this scenario, we followed RP-PCR classification and ignore the discrepant WGS classification.

### Compound Heterozygous Deleterious Variants near *RFC1* Gene

Raw FASTQ files of WGS data from the PNRR cohort were processed using Parabricks *pbrun germline* pipeline^32^ (version 4.0.0-1), which utilized GATK *HaplotypeCaller*^37^ as the variant calling tool. The resulting VCF files were split by chromosome and the following analysis were based on chromosome 4 only (where *RFC1* gene is located).

Variants in VCF files were annotated using snpEff^38^ (version 4.3) with multiple reference databases. After initial annotation against the Hg38 reference genome, the Ensembl^39^ GRCh38.86 database was used for gene annotation. Variant types were classified using SnpSift^40^, and the functional impact of variants on chromosome 4 was predicted with SIFT4G Annotator. This tool classifies amino acid substitutions as deleterious or tolerated based on sequence homology and physical properties of amino acids, with variants receiving a SIFT score < 0.05 considered potentially deleterious. As a result, no potential deleterious genetic variations were identified in the *RFC1* gene locus.

## Results

### WGS Study of *RFC1* STR Carriers

The workflow overview for this study is illustrated in **Figure 1**. Consecutively enrolled patients were selected from the PNRR cohort that were seen at the Johns Hopkins Hospital.^41^ A clinical diagnosis of iPN was established by examining physicians using a combination of symptoms and signs and exclusionary laboratory testing as defined by the American Academy of Neurology^42^. Only individuals meeting the criteria for iPN, defined as a slowly progressive, symmetric, distal axonal polyneuropathy of unknown cause, were selected for WGS. Patients with any other confirmed causes of distal symmetric peripheral neuropathies, such as amyloidosis, chronic renal failure, alcohol abuse, vitamin deficiencies, or inherited neuropathies (based on genetic diagnosis or neuropathy in a first-degree family member) were excluded, as were primary demyelinating neuropathies. The cohort consisted of 788 patients with iPN, each undergoing 30X Illumina short-read WGS. This cohort was 38.2% female and 92.9% European-ancestry individuals, with an average age at onset of 62.2 years (**Table 1**).

Using our WGS workflow, we performed case-control analysis and classified carriers of (AAGGG)_exp_ STR in the *RFC1* gene as either biallelic or monoallelic by applying unsupervised clustering methods on the percentage of reads containing the motif and the average detected STR length (**Figure 1** and **Methods**). Our pipeline identified significantly higher frequency of biallelic (AAGGG)_exp_ *RFC1* STR carriers in iPN patients (2.3%) than controls (0.1%; Fisher’s exact *p* = 0.00002, odds ratio [OR] = 19.9, 95% CI = [3.1 - 828.6]; **Figure 2A**). Similarly, we observed significantly higher monoallelic *RFC1* STR frequency in iPN (9.4%) compared to controls (6.3%; Fisher’s exact *p* = 0.02, OR = 1.5, 95% CI = [1.1 - 2.3]; **Figure 2B**). To address the absence of large-scale comprehensive benchmarking STR detection from short-read sequencing against RP-PCR, we performed PCR analysis on the same cohort analyzed by WGS.

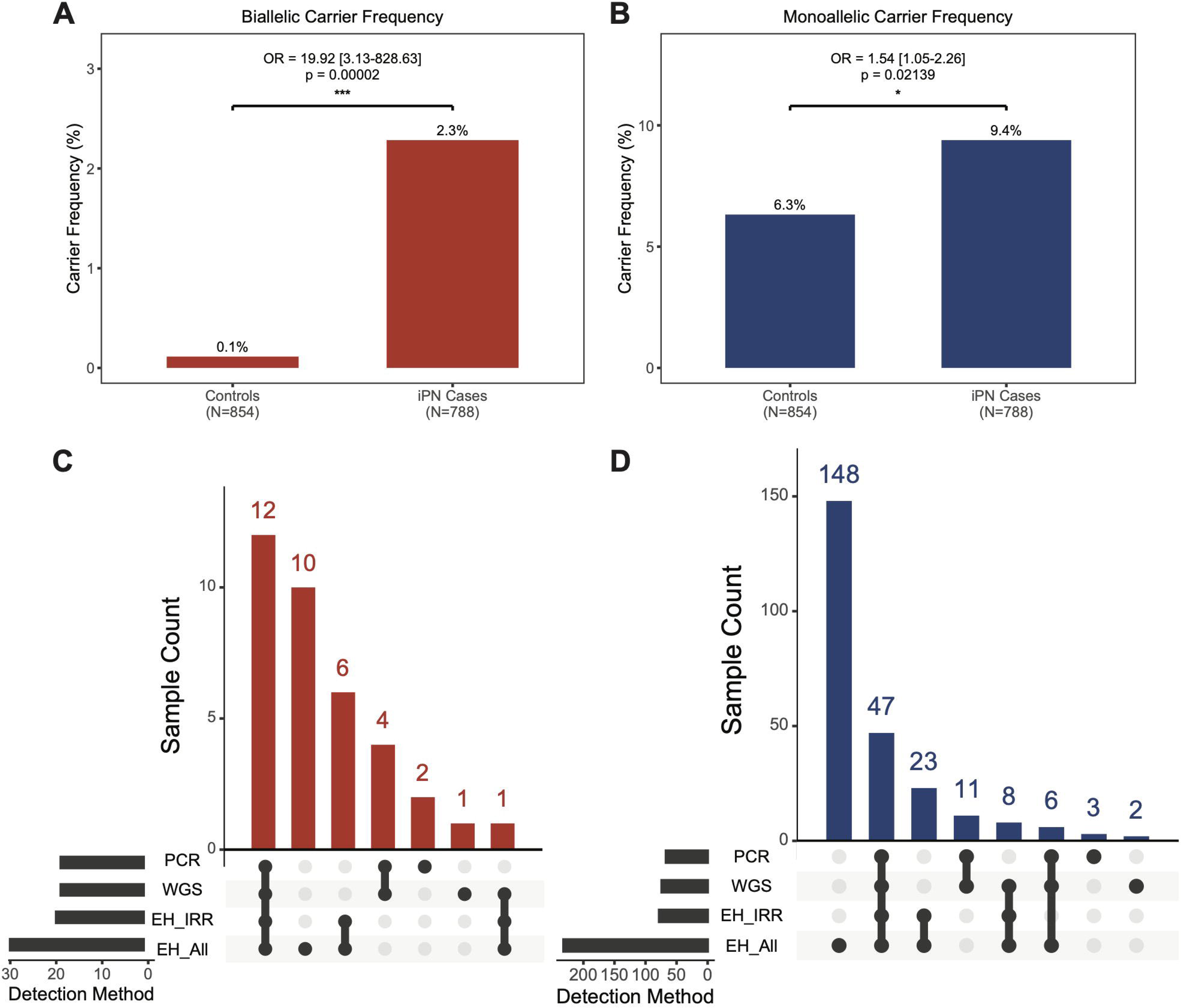

Our hybrid workflow, coupled with unsupervised clustering, demonstrated high precision in detecting STRs when compared to standard WGS analysis using EHdn or EH alone. As shown in **Figure 2C-D**, EH carriers with all types of supporting reads (last row in each plot) identified 10 biallelic and 148 monoallelic unique carriers that did not overlap with the shared biallelic and monoallelic carriers detected by PCR and our WGS workflow, suggesting a high rate of false positives. Restricting EH results to at least one allele having INREPEAT supporting reads (third row in each plot) helped eliminating some of these false carriers. Our workflow, building on top of EH INREPEAT calls and implementing our in-house genotyping and filtering criteria and further IGV examination, closely aligned with PCR results while maintaining a balance between sensitivity and specificity (see below).

### PCR Validation of *RFC1* STR Carriers

Next, we sought to validate WGS-based genotyping results by PCR. Overall, WGS and PCR methods produced matching genotype classifications in 98% (769/788) of iPN cases, with a Cohen’s kappa coefficient of 0.9, indicating excellent concordance (**Figure 3A**). Based on PCR validation result, we confirmed 10.2% (N = 80) of the iPN cohort as *RFC1* (AAGGG)_exp_ repeat carriers (**Figure 3B**). Among the carriers, 77.5% carried heterozygous repeat expansions (example shown in **Figure S2**) and 22.5% were biallelic repeat expansion carriers (example shown in **Figure S3**). In 5 samples that were classified as carriers by PCR only, we carefully reviewed the reads in IGV and found no reads carrying the altered AAGGG motif (example shown in **Figure S4**). We believe that this is due to the inherent alignment challenges in repetitive regions, where all AAGGG-containing reads likely mismapped to other genomic locations with similar repeat sequences. A detailed summary of the carrier status and genotyping results for all carriers is provided in **Table S2.**

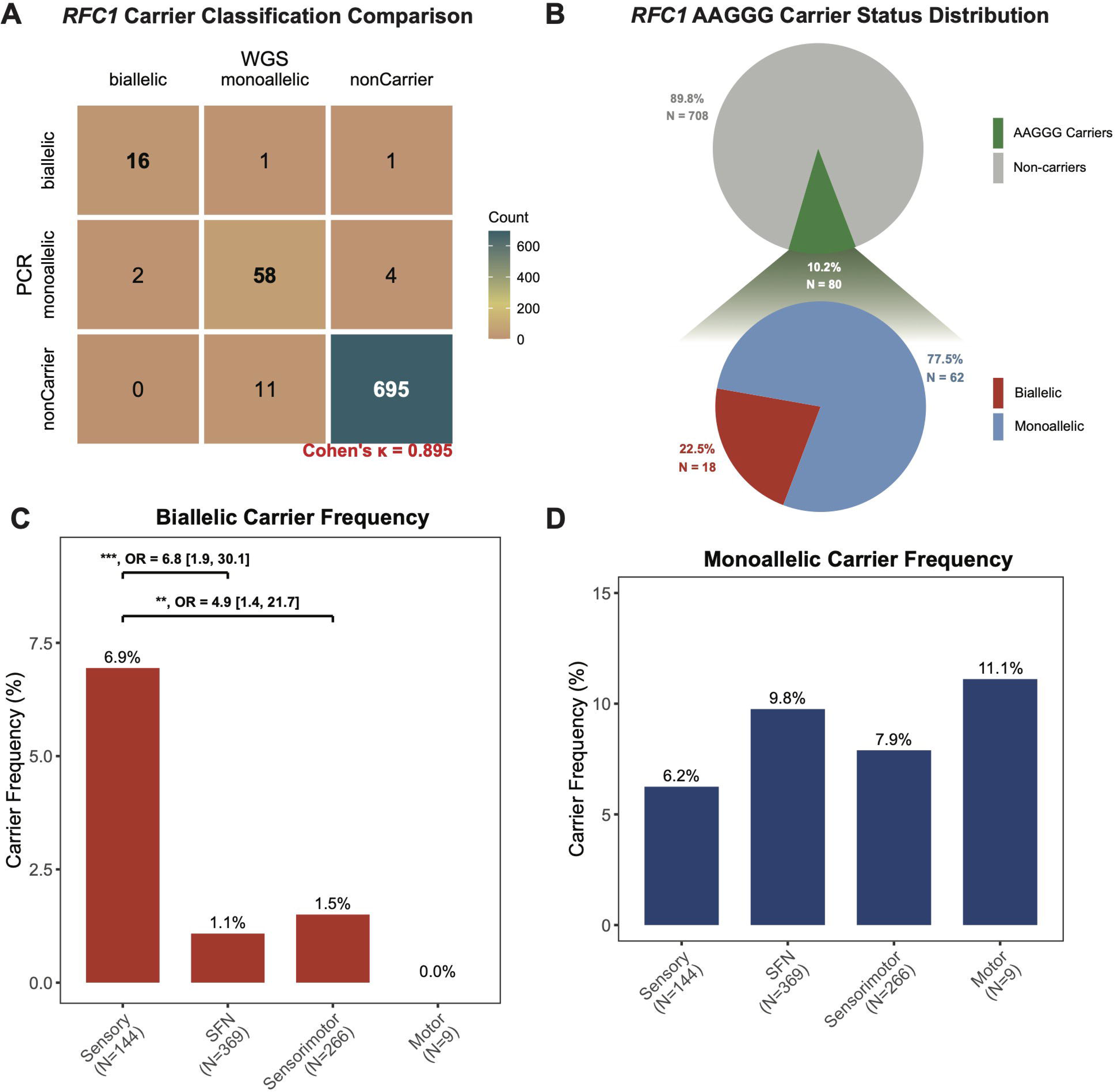

Stratified analysis based on PCR-validated carriers revealed that biallelic expansions carrier frequency was significantly higher in the pure sensory iPN subcategory (6.9%) compared to pure SFN patients (1.1%; Fisher’s exact *p* = 0.0008, OR = 6.8; 95% CI = [1.9 - 30.1]) and sensorimotor IPN patients (1.5%; Fisher’s exact *p* = 0.008, OR = 4.9; 95% CI = [1.4 - 21.7]; **Figure 3C**). No significant differences in biallelic carrier frequency were observed between pure SFN and sensorimotor iPN subcategories or between sensorimotor iPN patients and pure motor iPN patients. The elevated biallelic carrier frequency in the sensory subtype aligned with previous reports, but the frequency of biallelic repeats in this population (6.9%) was lower than previously described (34%).^21,43^ In contrast, a stratified analysis on monoallelic (AAGGG)_exp_ *RFC1* STR carriers revealed no significant differences between sensory, SFN, sensorimotor, and sensory/SFN categories (**Figure 3D**).

### PCR Validation of Atypical Repeat Carriers

We saw several different types of atypical repeat carriers in our analysis. For instance, we observed four samples with stretches of AAGGG mixing with other repeat motifs on both alleles. To differentiate them from true AAGGG expansion carriers, we performed additional motif-specific RP-PCR and fragment analysis for motifs AAAGG, ACAAG, AAAGGG, AAGAG, and AACGG. In these samples, all motifs (including AAGGG) showed peaks rather than the typical decremental sawtooth pattern in fragment analysis, exhibiting no definitive evidence of true AAGGG expansions (**Figure 4**). Therefore, we excluded these samples from the AAGGG expansion carrier list in our analysis.

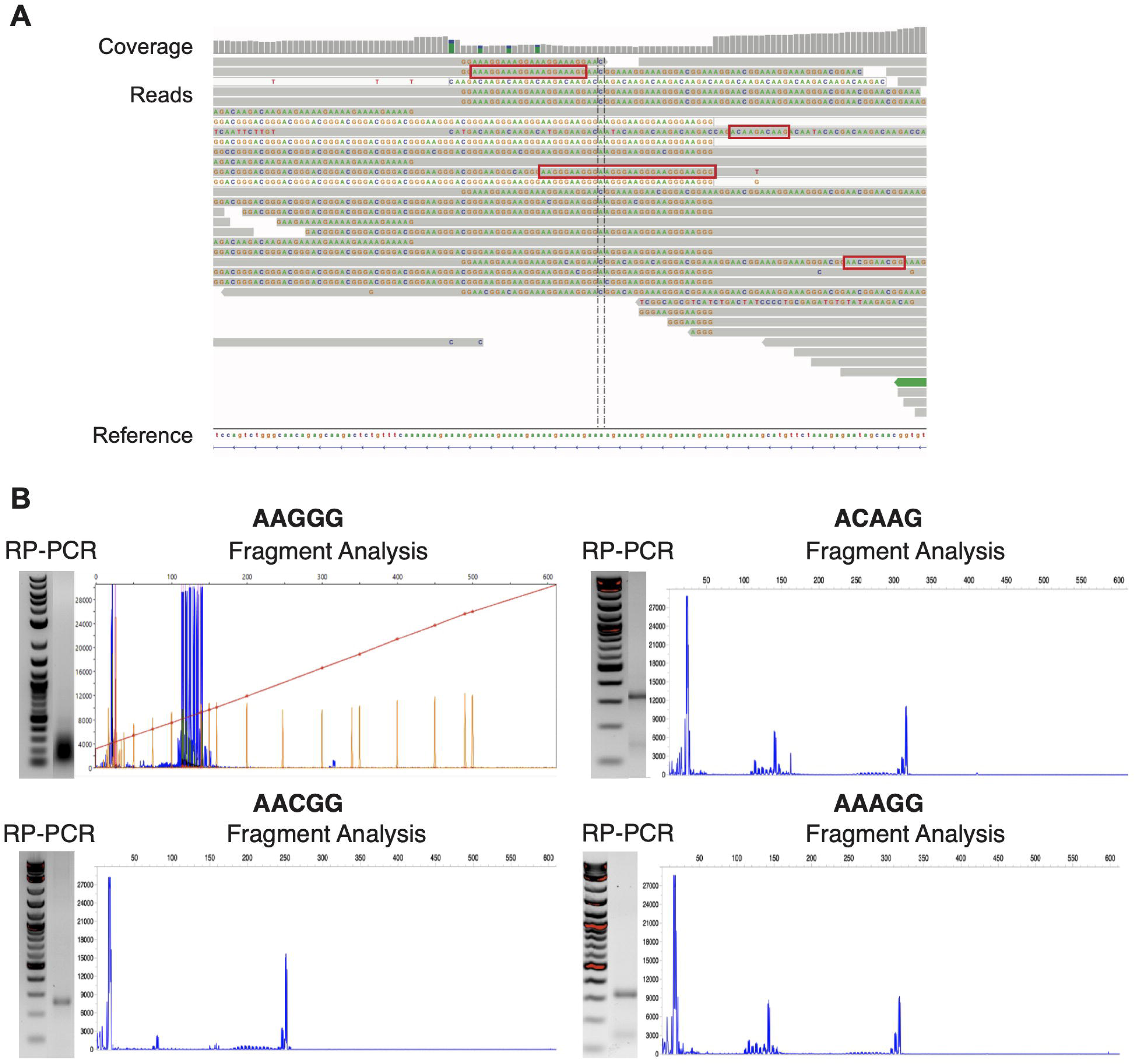

In addition, we also observed five patients carrying *RFC1* expansions with other motifs on one or more alleles (**Table S4**). Although these alternative *RFC1* motifs were rare in our cohort, a recent long-read sequencing study reported their presence in self-reported Black or African American participants from the All of Us program.^44^ Given the high prevalence of peripheral neuropathy in the general population, it is possible that these novel *RFC1* motifs contribute to disease risk, a question that warrants further investigation.

### Clinical Spectrum of Patients Carrying *RFC1* STRs

We next analyzed the clinical characteristics of the confirmed 18 biallelic and 62 monoallelic *RFC1* (AAGGG)exp in comparison to 708 non-carriers (**Figure 3B**; **Table 2**). A two-way chi-squared test (degrees of freedom DOG = 2) revealed a significant difference in the prevalence of sensory iPN (*p* < 0.001), with biallelic carriers showing markedly higher rates (56%) than monoallelic carriers (13%) or non-carriers (18%). No significant differences were observed across the three groups with respect to sex, age, presence of severe pain, sensorimotor iPN, pure SFN, or motor iPN.

Given that AAGGG repeat expansions classically cause CANVAS, which features ataxia and vestibular features, we looked for additional clinical features which might capture subclinical phenotypes that might suggest an *RFC1* expansion when evaluating a patient with idiopathic neuropathy. A previous study reported no association between age at onset and the number of AAGGG repeat units in biallelic carriers.^20^ In our study, the average age of onset in biallelic carriers and monoallelic carriers was mid 50s, lower than the average age of the entire cohort (62.2 years old). Several of the repeat positive biallelic SFN patients had a relatively acute onset neuropathy in their 20s and 30s. The most common clinical features in our cohort included distal sensory loss and difficulty in walking due to balance issues.

## Discussion

This study represents the largest genetic investigation to date of a single, consistently and thoroughly phenotyped iPN cohort. It is also the first comprehensive comparative analysis of two complementary molecular approaches: WGS and RP-PCR. We developed an integrated pipeline that utilizes EHdn and EH algorithms for *RFC1* STR detection from short-read WGS, achieving 98.2% agreement with PCR-based validation results. Additionally, this pipeline incorporates unsupervised clustering methods to accurately classify patients as biallelic or monoallelic *RFC1* AAGGG expansion carriers.

Using this pipeline, we observed that biallelic (AAGGG)_exp_ in *RFC1* accounted for 2.3% of iPN cases and was detected in only one control subject. The frequency of biallelic carriers was significantly higher among patients with pure sensory iPN (6.9%). However, compared to prior findings by Curro et al.^19^, we observed a much lower overall rate of biallelic expansions in pure sensory iPN (6.9% versus 34%). The lower overall rate of biallelic expansions in iPN we observe is largely consistent with unpublished clinical observations of several large subspecialty clinics focused on peripheral neuropathy. The reason for this discrepancy is unclear. Previous studies focused primarily on patients from England or Italy, and expansions do appear more prevalent in European populations.^45^ However, the PNRR cohort is heavily biased towards Caucasians and thus likely reflects a genetic background similar to prior European studies. Differences in selection and screening criteria and repeat assessment methodology as well as the smaller size of the Italian cohort, which included only 125 cases, may also explain the discrepancy.^19^ Similarly, identification of biallelic *RFC1* repeat expansions in SFN and sensorimotor iPN patients in our large cohort differs from observations in smaller and ataxia focused cohorts, where a prior study^19^ reported no biallelic carriers among 100 patients with sensorimotor iPN. In contrast, our larger cohort revealed biallelic *RFC1* expansions in 1.5% of such patients. Thus, we suggest that the presence of motor nerve involvement should not preclude patients from undergoing *RFC1* repeat screening.

Interestingly, monoallelic *RFC1* (AAGGG)_exp_ showed a significantly higher prevalence in our iPN cohort (9.4%) compared to controls (6.3%; *p* = 0.02), suggesting that a single expanded *RFC1* allele may confer an increased risk for developing iPN, though the mechanism and penetrance may differ from the established pathogenic role of biallelic expansions. Consistent with this notion, the monoallelic prevalence observed in iPN cases also exceeded the 7.3% reported by Kristina et al. in a large multi-ethnic disease cohort from TOPMed and the 100K Genomes Project.^45^ Prior studies suggest that some CANVAS cases have compound heterozygous mutations—with a AAGGG repeat on one allele and a protein-altering mutation on the other allele. However, we observed no such potential damaging missense, loss-of-function variants, or small insertions/deletions near the *RFC1* region in our monoallelic *RFC1* carriers or in our controls (**Methods**). We did observe short (typically 2-5 copies) alternative repeat sequence elements within the other allele in 19 out of 62 (30.6%) of our monoallelic carriers that might provide an alternative explanation for the increased frequency in the iPN population. However, all such cases had AAAAG repeat alleles as measured by repeat-primed PCR and thus met our criteria for monoallelic carrier status.

Our control cohort included rare *RFC1* AAGGG biallelic carriers (0.1%), which was similar to the 0.15% reported in a previous short-read WGS study^45^. Likewise, monoallelic carriers in our controls occurred at a frequency of 6.3%, comparable to the 7.3% reported in TOPMed^45^ and higher than the 0.7% reported in the 1000 Genomes project^46^. Interestingly, we observed distinct control carrier frequencies when we analyzed *RFC1* STRs using different EH catalogs, including the EHdn-defined region (chr4:39,348,426-39,349,038), the RepeatMasker–defined region (chr4:39,348,275–39,348,633), and coordinates from Ibanez et al. ^45^ (chr4:39,348,424–39,348,479; **Figure S5**). This finding highlighted the technical sensitivity of STR detection algorithms, where using a larger region-of-interest, as defined in EHdn catalog, led to a significantly lower carrier frequency compared to other catalogs used. A limitation of this case-control study is that some of our control subjects (who were recruited as controls for an Alzheimer study) may have had peripheral neuropathy based on reported symptoms. As such, our numbers may under-estimate the impact of monoallelic *RFC1* (AAGGG)_exp_ on the risk of developing iPN. This factor will need to be carefully considered in future large database analyses.

This finding of enriched monoallelic cases in iPN has implications for our understanding of how non-reference AAGGG repeat expansions might elicit disease. If the repeat elicits disease through a loss of function mechanism, this suggests that even haploinsufficiency for *RFC1* may enhance the risk of developing neuropathy. Alternatively, heterozygous AAGGG repeat expansions may induce a dose-dependent gain of function toxicity that would induce more fulminant symptoms and greater penetrance in homozygosity.^29^ This latter model would be consistent with recent studies in CANVAS patient derived glutamatergic iNeurons, where selective removal of the repeat suppressed disease relevant phenotypes, but reprovision of RFC1 protein did not.^29^ Future work will be needed to more clearly ascertain the impact of heterozygous mutations on peripheral sensory neuronal dysfunction and disease pathogenesis.

There are several cases in our cohort of monoallelic *RFC1* repeat expansion in young individuals with relatively acute onset SFN. Although the presentation of these patients resembled monophasic acute-onset post-infectious SFN, they lacked a history of prior infection, and their neuropathy persisted for many years. This finding suggests that acute onset SFN patients who do not recover in a timely manner should be evaluated for *RFC1* repeat expansions. It also raises the possibility that monoallelic expansions may serve as a risk factor for poor recovery from peripheral nerve injury or neurotoxic agents.

Comparison between short-read WGS with PCR revealed excellent concordance for most cases. Aside from AAGGG repeat carriers, we found some carriers with other repeat structures, such as (AAAGGG)exp, (AAAGG)exp, or a mixture of them with (AAGGG)exp (**Table S4**). These cases were initially identified by WGS and subsequently validated by RP-PCR. Our WGS pipeline successfully identified these alternate repeat motifs that would likely have been overlooked by conventional PCR assays designed for a limited set of target sequences, demonstrating the superior sensitivity of WGS for comprehensive variant screening. Our automated pipeline offers significant practical advantages for clinical implementation. The in-house pipeline, written in Python, Bash, and R, can be easily implemented in clinical settings by personnel with basic bioinformatic training. While long-read sequencing offers a more definitive characterization of STR expansions, its cost and technical requirements (e.g., large amounts of gDNA) currently limit routine use. Our approach provides a practical, cost-effective alternative for detecting *RFC1* expansions, with WGS complementing PCR by flagging cases for targeted follow-up validation. We anticipate this tool will be broadly adopted for both research and clinical applications, facilitating more accurate and efficient genetic analyses in the field of human and clinical genetics. By making our pipeline publicly available through the cloud-based platform (GitHub), we aim to facilitate broader adoption of this methodology in both research and clinical settings, providing additional diagnostic yield and deepening the understanding of the genetic basis of PN. Ultimately, we hope to facilitate more accurate and efficient genetic analyses in the field of human and medical genetics.

This study has some limitations. Both WGS and PCR methods face challenges in precisely quantifying repeat number, particularly in the presence of multiple repeat motifs. Classifying carriers can be particularly challenging when reads are interrupted by various motif combinations, since capture success for both PCR and WGS depends on motif composition and regional complexity. While long-read sequencing would address these shortcomings, it requires significantly larger DNA input and remains prohibitively expensive for large-scale use and is therefore best reserved for cases where WGS and PCR cannot resolve allelic information or when precise measurement of repeat length is required.

Additionally, the limited number of biallelic *RFC1* carriers identified in this study precludes definitive conclusions regarding the spectrum of affected fiber types and optimal patient selection for screening. While our findings suggest that *RFC1* repeat expansion can occur across the full spectrum of iPN patients, including pure small fiber and motor presentations, we cannot firmly determine whether specific clinical features predict *RFC1* expansion carrier status. Larger-scale studies with increased numbers of confirmed biallelic expansion carriers will be necessary to establish whether *RFC1* screening should be uniformly applied to all iPN patients or targeted to specific phenotypic subgroups.

In conclusion, our findings underscore the diagnostic value of *RFC1* expansion screening in iPN patients, where biallelic expansions account for roughly 2.3% of cases, and monoallelic expansions occur at a significantly higher frequency in iPN patients versus controls. Overall, our data support testing for *RFC1* repeat expansions in patients with iPN. Moreover, our finding that monoallelic expansions may serve as a risk allele for the development of peripheral neuropathy has important implications for how we counsel our patients both before and after testing. Considering the current difficulties in obtaining *RFC1* testing in the United States, our pipeline will be very useful as a rapid screening test for *RFC1* repeat expansion carriers. We are hopeful that applying this pipeline can lead us to a deeper understanding of genotype-phenotype relationships will impact our ability to resolve the pathogenic mechanisms underlying CANVAS and peripheral neuropathy as a whole – providing a way forward for developing therapies for these currently untreatable conditions.

## Data Availability

All raw genomic data supporting the findings of this study have been deposited in the Database
of Genotypes and Phenotypes (dbGaP) under the accession number phs003788.v1.p1.

## Acknowledgements

We would like to thank the patients, their families, and the Foundation of Peripheral Neuropathy for their support of the PNRR and their willingness to participate in research. We appreciate obtaining access to genetic and phenotypic data from the participants. We also thank the PNRR Study Group members at Johns Hopkins who enroll patients in the registry. These include neuromuscular physicians Sarah Berth, Vinay Chaudhry, David Cornblath, Leana Doherty, Lindsey Hayes, Hristelina Ilieva, Thomas Lloyd, Mohammad Khoshnoodi, Brett McCray, Brett Morrison, Bipasha Mukherjee-Clavin, Lyle Ostrow, Michael Polydefkis, Ricardo Roda, and Charlotte Sumner.

We thank the Genome Technology Access Center at the McDonnell Genome Institute at Washington University in St. Louis for DNA sequencing.

We are grateful to all of the families participating in the Undiagnosed Mendelian Disorders (UMD) Research Project at Washington University in St. Louis, as well as the principal investigator and team members of the UMD project (M. Shinawi, D. Baldridge, C. Gurnett, T. Turner, B. Leon Ricardo). We appreciate obtaining access to genetic and phenotypic data from the UMD project.

PNRR is supported by the Foundation for Peripheral Neuropathy. SCJ is supported by the Hydrocephalus Association Innovator Award, Cerebral Palsy Alliance Research Foundation Project Grant (PRG03121), WashU Children’s Discovery Institute Faculty Scholar Award (CDI-FR-2021-926), and NIH U19NS130607, R01NS111029, and R01NS131610. AH is supported by NIH (R21NS135481, P30 MH075673-011), Wellcome Trust, Dr. Miriam and Sheldon G. Adelson Medical Research Foundation, and Dr. Richard Merkin Family Foundation. JM is supported by the NIH U19NS130607 and the Bob and Signa Hermann Fund for Neuropathy Research. PKT and SSO’s work on this project was supported by the NIH (R21NS129096) and the Taubman Institute at the University of Michigan.

## Author Contributions

Z.T., S.C.J., P.K.T., A.H., J.M., S.S.O. contributed to the conception and design of the study; Z.T., S.S.O., R.I., S.C.J., P.K.T., A.H., J.M., B.C., E.M., S.T., J.U., Z.L., D.B., C.C., M.J. contributed to the acquisition and analysis of data; Z.T., S.C.J., S.S.O. contributed to drafting the text or preparing the figures and tables. All authors contributed to editing the manuscript.

