## Supplementary figures and images for "Heterozygous and Homozygous *RFC1 AAGGG* Repeat Expansions are Common in Idiopathic Peripheral Neuropathy"

### Figure S1

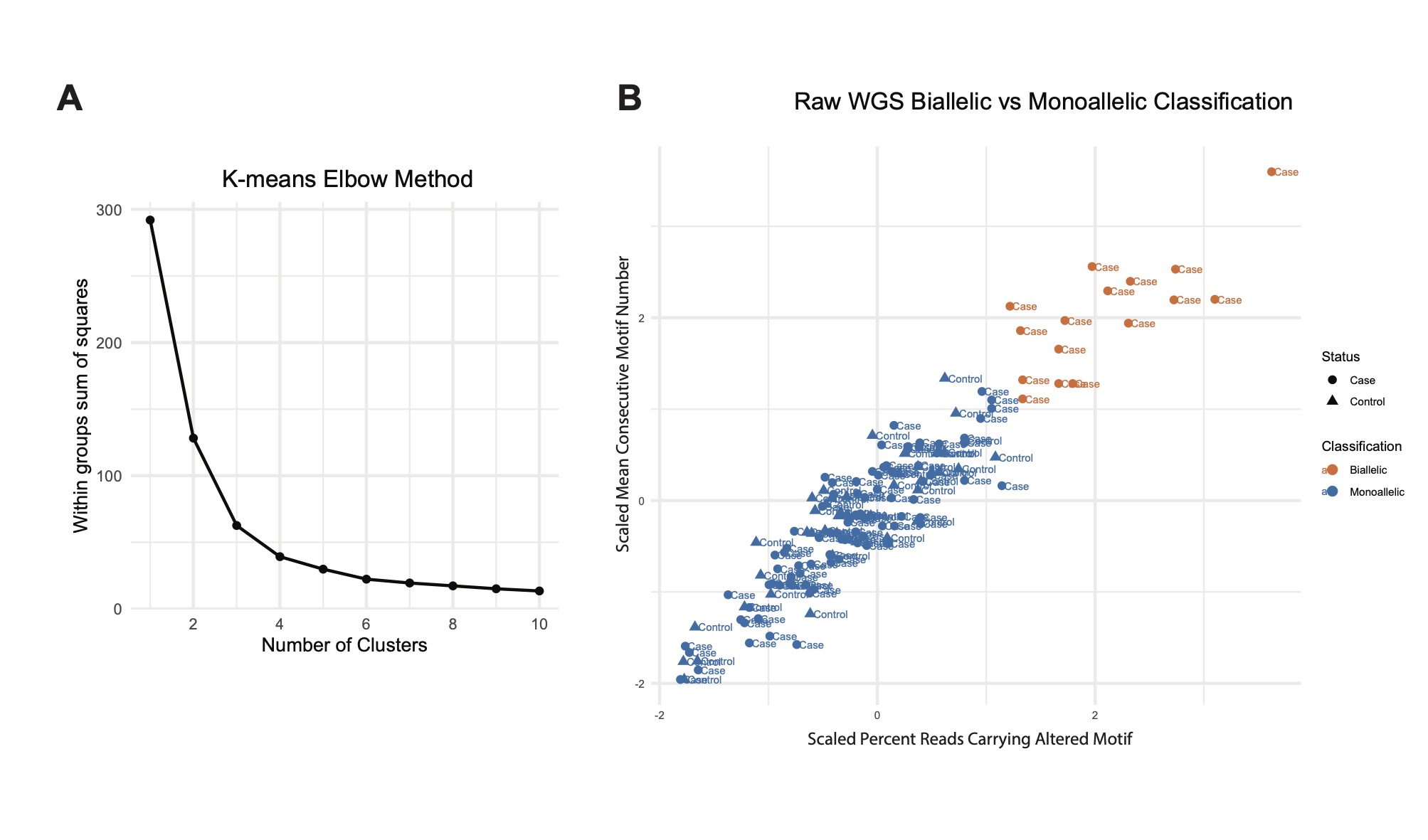

### Figure S2

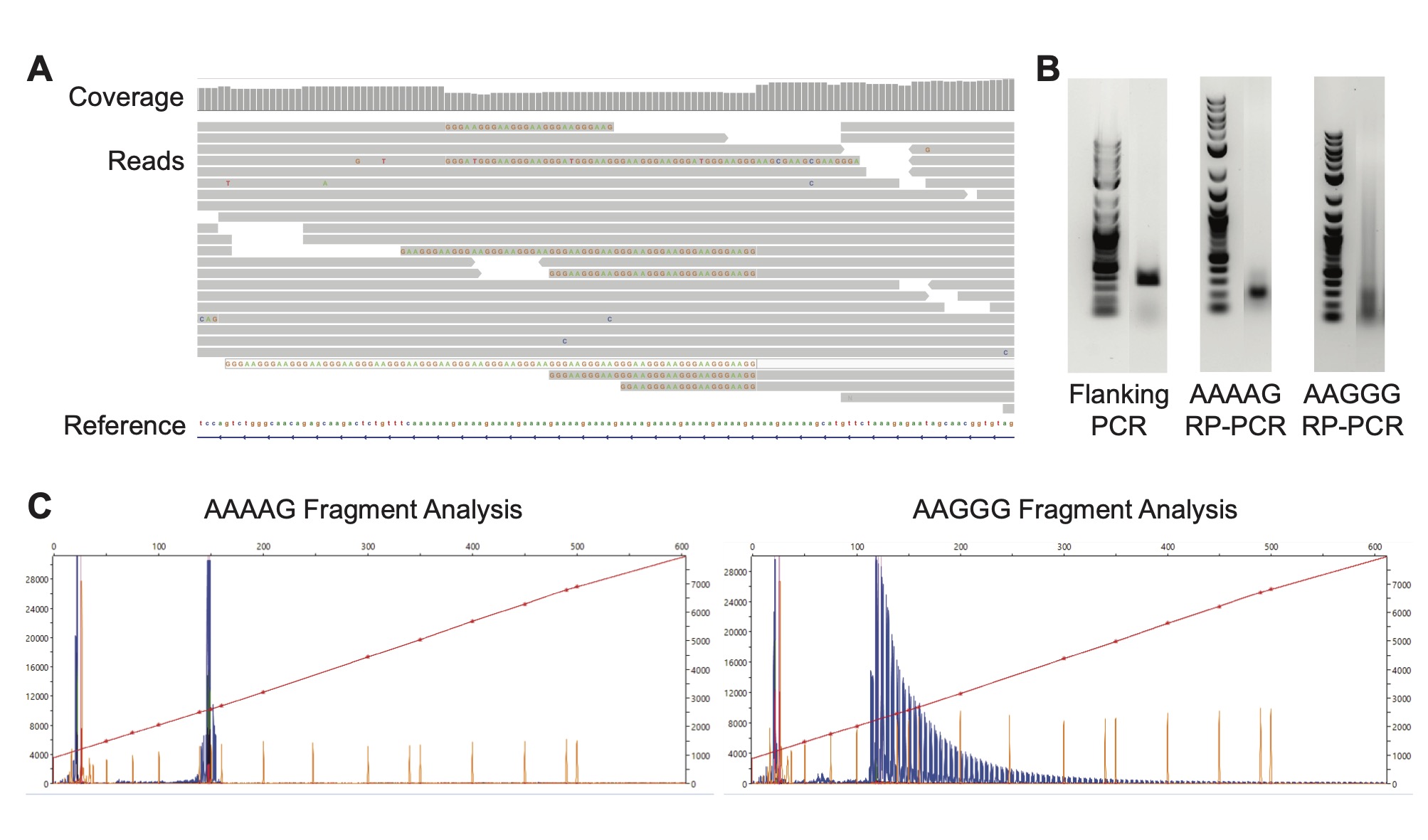

### Figure S3

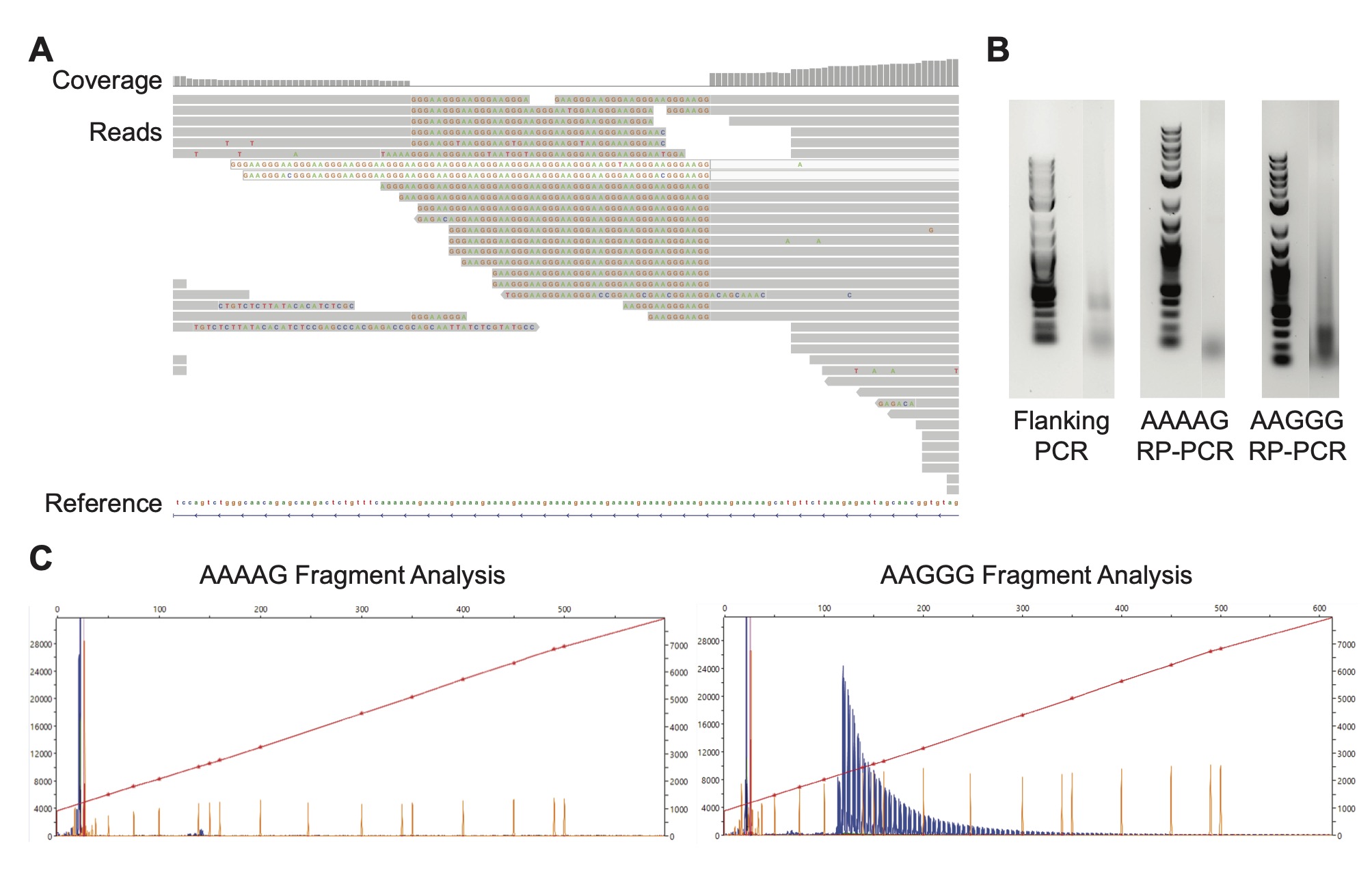

### Figure S4

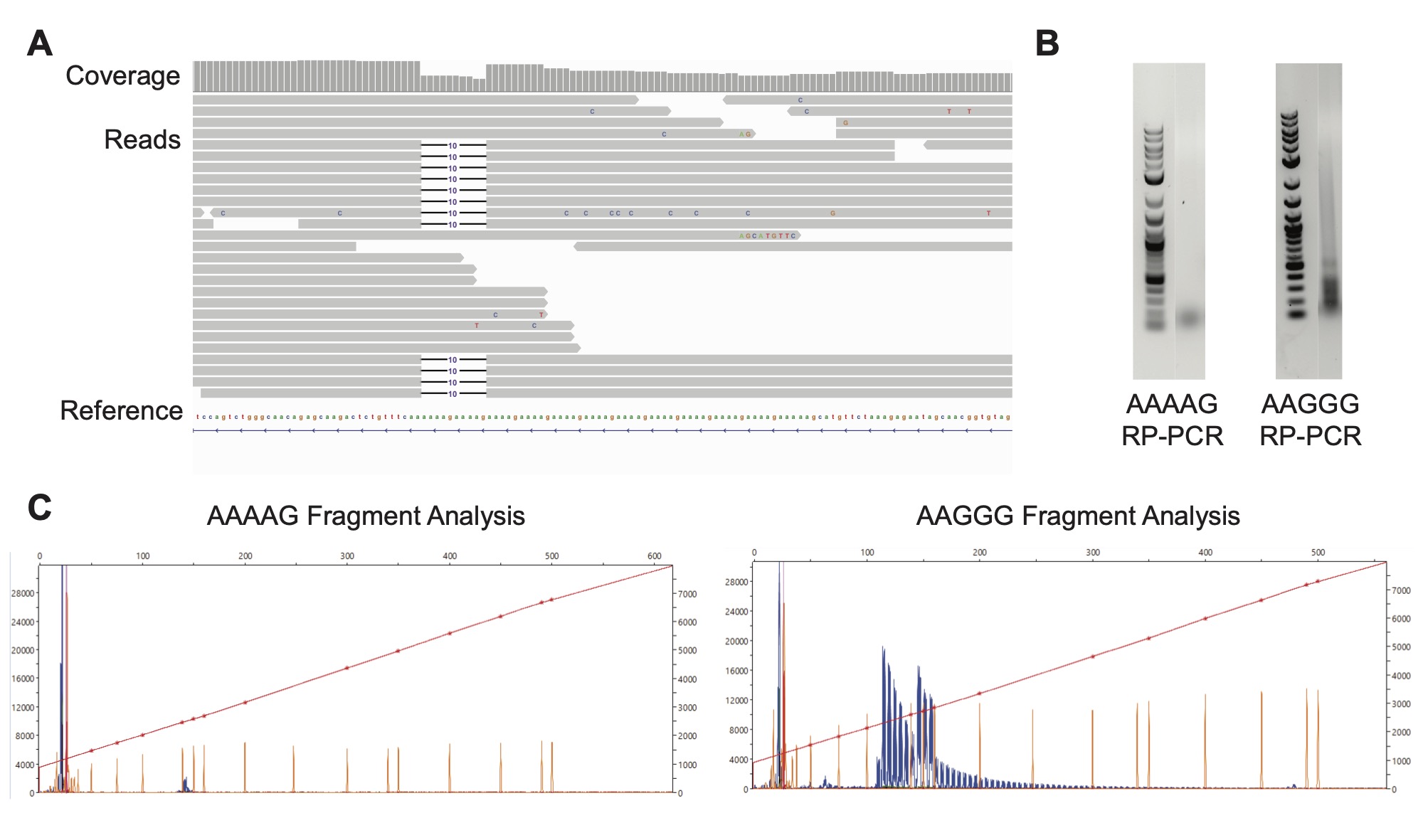

### Figure S5

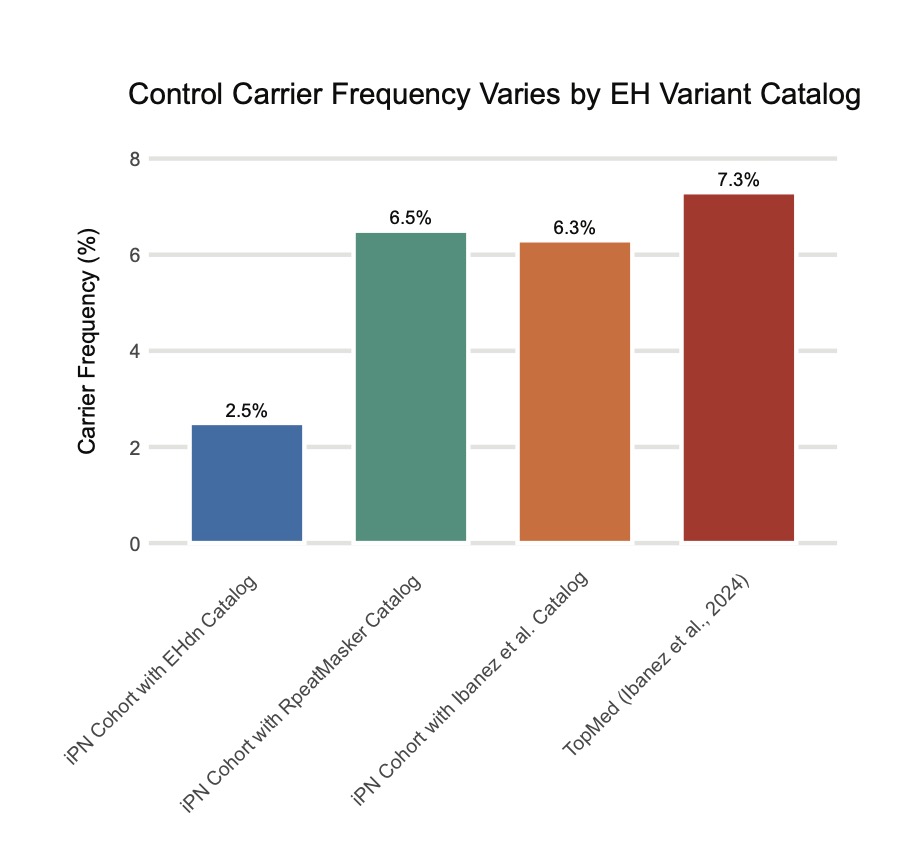
